# Silica-encapsulated DNA tracers for measuring aerosol distribution dynamics in real-world settings

**DOI:** 10.1101/2021.05.19.21257392

**Authors:** Anne M. Luescher, Julian Koch, Wendelin J. Stark, Robert N. Grass

## Abstract

Aerosolized particles play a significant role in human health and environmental risk management. The global importance of aerosol-related hazards, such as the circulation of pathogens and high levels of air pollutants, have led to a surging demand for suitable surrogate tracers to investigate the complex dynamics of airborne particles in real-world scenarios. In this study, we propose a novel approach using silica particles with encapsulated DNA (SPED) as a tracing agent for measuring aerosol distribution indoors. In a series of experiments with a portable setup, SPED were successfully aerosolized, re-captured and quantified using quantitative polymerase chain reaction (qPCR). Position-dependency and ventilation effects within a confined space could be shown in a quantitative fashion achieving detection limits below 0.1 ng particles per m^3^ of sampled air. In conclusion, SPED show promise for a flexible, cost-effective and low-impact characterization of aerosol dynamics in a wide range of settings.

**PRACTICAL IMPLICATIONS:** For the first time, silica particles with encapsulated DNA were used to characterize a confined indoor space regarding position- and ventilation-dependent effects of aerosol distribution. The method described here introduces SPED as a novel, non-toxic, low-impact, cost-effective and easy-to-use aerosol tracing platform that can be used to examine real-world environments. The mobile setup presented here as a proof of concept shows that SPED can be aerosolized and re-captured, followed by highly sensitive quantitative barcode-specific PCR analysis. The results revealed that this tracing method can detect position-dependent differences in exposure and ventilation effects influencing distribution dynamics. In the future, SPED could be engineered to exhibit custom-designed properties and be employed within a wide range of setups and high-capacity multi-tracing combinations.

## INTRODUCTION

Aerosol dynamics are an important factor when assessing the circulation of hazardous pollutants and pathogens with regard to human health and the environment. Viable bioaerosols are a well-known cause for many infectious diseases such as tuberculosis^1^, measles^2^, Legionnaire’s disease^3^, influenza^4^, gastroenteritis^5^ and SARS–CoV-1^6^ and SARS-CoV-2^7–10^. With the COVID-19 pandemic spreading globally, the importance of understanding the mechanisms of aerosol spreading is perhaps more evident than ever and discussions surrounding the topic have reached the mainstream. Specifically, the assessment and characterization of aerosol distribution properties within specific indoor spaces such as lecture halls, office spaces, hospitals, public transport vehicles or event venues is of high interest with regard to public and occupational health.

Many of the existing approaches to assess indoor environments regarding aerosol dynamics rely on computational models. However, such models are based on the pre-existing understanding of fluid dynamics and prone to numerical error^11^, while also depending on data for setting realistic parameters^12^. Therefore, predicting real-world behavior in complex, non-controlled environments is computationally challenging. For those reasons, physical tracing is still a vital field to assess aerosol-related properties in real-world settings to help validate and complement model-based studies.

Table 1 shows a list of studies reporting tracing methods to characterize (bio-)aerosol distribution indoors. Carbon dioxide, for example, is a commonly used tracer gas, applied either alone or in combination with other tracers, and in conjunction with one or more CO_2_ detectors. For example, Knibbs et al.^13^ employed CO_2_ to assess the effect of ventilation rates in various room settings, using the results to model virus-specific infection risks. While CO_2_ is widely accessible, non-toxic (in low concentrations), easily detected and self-clearing, it is also naturally present in ambient air. Subtle effects may therefore be harder to detect and relatively high working concentrations are needed. Moreover, aerosol dynamics are complex and not necessarily approximated accurately by a non-particulate tracer gas.

**Table 1.** Literature overview listing aerosol tracing methods for room characterization

| Method | Tracer type | Tracer size | Means of aerosol generation/distribution | Means of collection | Method of quantification | Time-/location-resolved results | Lower limit of detection | Specificity for tracer | Tracer occurring naturally in environment | Toxicity/infectiousness at working conc. | Multi-tracing possible |
| --- | --- | --- | --- | --- | --- | --- | --- | --- | --- | --- | --- |
| Schade et al.<sup>14</sup> | DEHS<sup>a</sup> (in conjunction with CO<sub>2</sub>) | 300 nm (maximum of size distribution) | Aerosol generator with integrated pump | Handheld particle counters | Optical light scattering (particle counter) | Yes/yes | 1 µg/m<sup>3</sup> resolution in time-resolved measurement<sup>b</sup> | No | No (DEHS)/yes (CO<sub>2</sub>) | No | No |
| Pyankov et al.<sup>15</sup> | Influenza A virus | n/a | 3-jet Collison nebulizer | Bioaerosol sampler | PCR | No/ | n/a | Yes | Yes | Yes | No |
| LUQAS study<sup>16</sup> | Artificial saliva | n/a | Particle generator<sup>c</sup> | No collection | LDA/PDA and PIV | Yes/yes | n/a | No | No | No | No |
| Van Rijn et al.<sup>17</sup> | Glycerol/ethanol | Same size distribution as cough droplets | Specially designed spray nozzle | No collection | SprayScan® laser sheet | Yes/yes | n/a | No | No | No | No |
| Knibbs et al. | CO<sub>2</sub> | No droplets | No aerosols | No collection | CA-10 CO<sub>2</sub> analyzer | Yes/no | n/a | Yes | Yes | No | No |
| This work | Silica-encapsulated DNA | Ca. 150 nm<sup>d</sup> | Airbrush gun | BioSampler Impinger | PCR | Yes/yes | < 0.1 ng/m<sup>3</sup> | Yes | No | No | Yes |

a. Di-ethylhexyl-sebacate.
b. For the devices used in the study, no lower limit of detection is available. For the Fidas® Frog, used for position-dependent counting, a measuring range of 0-20'000'000 p/L is indicated by the manufacturer. For the PCE devices used for time-resolved sampling, the resolution is 1 µg/m<sup>3</sup> according to the product information, meaning the limit of detection would be equal to that, or higher.
c. Not further specified, but image provided in study
d. Mean of hydrodynamic size distribution of tracer agent in suspension.

Schade et al.^14^ therefore combined CO_2_ with an aerosol tracer to characterize distribution in a large concert hall. Their combinatorial approach has the advantage of monitoring aerosol droplets with temporal and spatial resolution, with the limitation that the particle counters used for detection are not specific to the tracer aerosols and the resolution of detection is only in the µg/m^3^ range.

Another approach, by Pyankov et al.^15^, uses Influenza A virus as a tracer and a personal sampler for detection. However, they focus on introducing a new method of detection rather than a routine tracer entity for room characterization. Any approach using live viruses is not routinely feasible in a real-world setting as it requires high safety precautions, well-trained personnel and extended experimental preparation time.

Further physical tracers^18–24^, typically used outdoors and for environmental tracing, exist, but they, too, face challenges related to either environmental background concentrations, hazard and toxicity or non-specificity of the detection method.

None of the reviewed tracing methods are suitable for complex multi-tracing or offer an extensive platform for flexible tuning of properties. There is still an unmet need for additional tracing methods allowing for direct and reliable experimental real-world characterization of indoor spaces regarding (bio-)aerosol distribution. Hence, novel tracing materials that are cost-effective, non-toxic and can be detected with a high sensitivity and specificity in a broad range of scenarios would be a valuable addition to existing tools.

We propose a new approach using aerosolized Silica Particles with Encapsulated DNA (SPED) to analyze aerosol dynamics. These sub-micron particles stand out in their ease of synthesis, durability and quantitative analysis with a low detection limit, as described by Paunescu et al.^25^ Silica is recognized as “safe” by the FDA with silica nanoparticles being used as food additive and in medicine (e.g. imaging).^26^ The encapsulated artificial DNA, or “DNA barcode”, is not present in the environment and can be detected with an extraordinarily high selectivity and sensitivity using real-time quantitative polymerase chain reaction (qPCR). This powerful detection method is known for its use in diagnostics to identify various pathogens, for example in food^27^, water^28^ and also aerosols^15, 29, 30^. Likewise, SPED are already established as tracers for liquids and surfaces and have been used to characterize aquifers^31^, assess pesticide drift^32^, track and trace comodities^33, 34^ and observe trophic interactions within food webs.^35^ Moreover, they have been employed as surrogate tracers for bacteria in a hospital environment^36^. Similarly, bacteriophages, *C. difficile* spores and cauliflower mosaic DNA have been employed for surface tracing in various settings, sometimes in conjunction or compared with fluorescent markers^37–42^. The DNA sequences are unique identifiers, offering a great advantage over other methods by allowing for specific detection. In the case of SPED, the sequences are synthetic, not bound to an organism, can be changed at will and are more robust through silica encapsulation. Since even a sequence length of just 60-100 nucleotides offers trillions of potential barcodes, a system with nearly unlimited multi-tracing capabilities is conceivable. Thus, the advantages of highly sensitive PCR detection can be implemented without the need for tracing organisms.

In this study, SPED are dispersed as aerosols and re-collected using commercial biosampler impingers, which offer a proven method to capture aerosols^43–45^. The DNA is released from its silica protection and quantitatively analyzed. The overall principle is summarized in Figure 1. Two batches of SPED, S1 and S2, were used, which differ in the sequence of their DNA barcode. Working with these unique identifiers has the advantage that they enable a wide variety of experiments, largely eliminating contamination between experiments and permitting scenarios such as simultaneous multi-source sampling.

**Figure 1:**
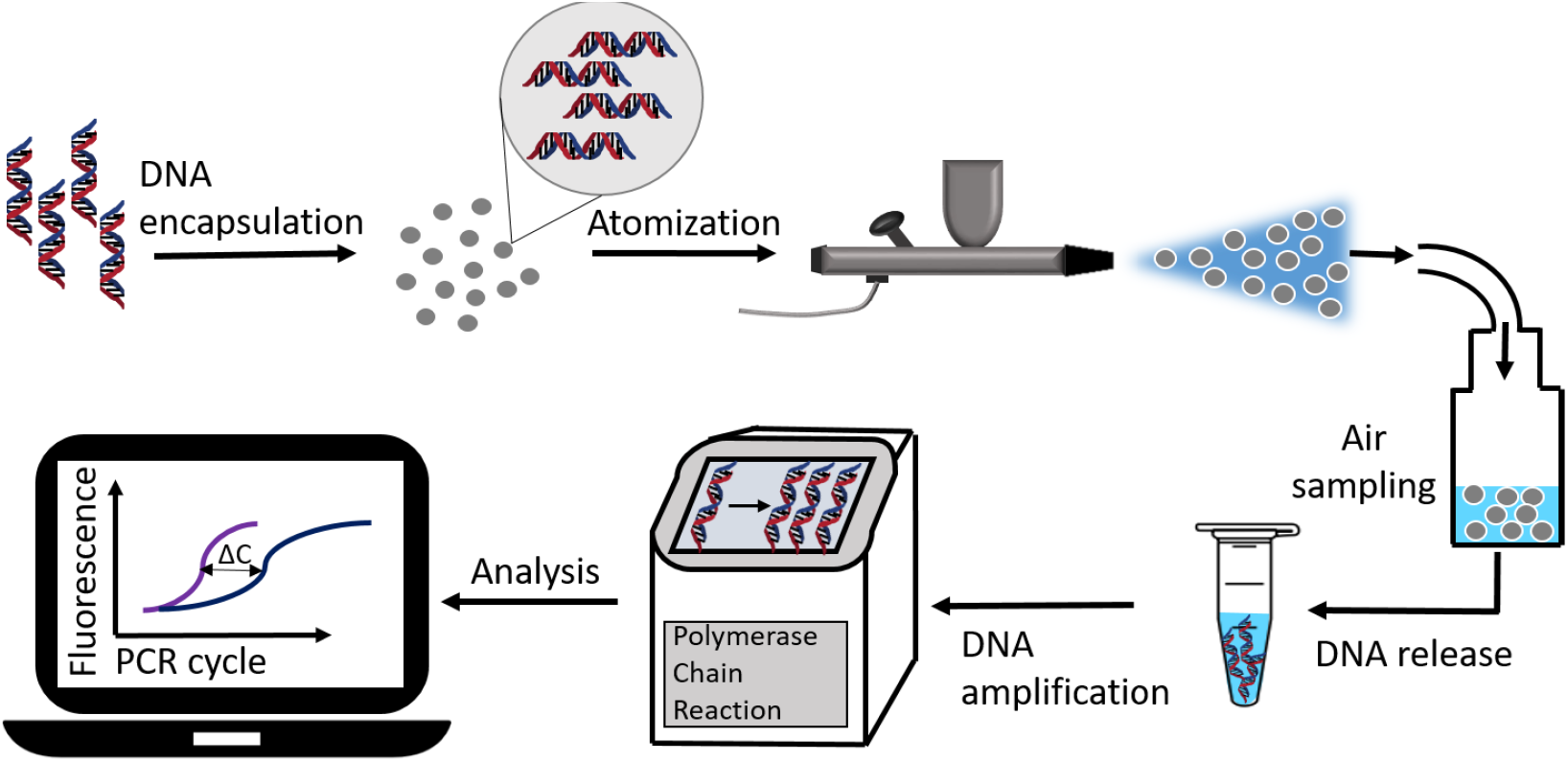
Schematic description of the tracing method using SPED. DNA is encapsulated in silica particles to form SPED, which are then aerosolized using an airbrush gun. The air is sampled using glass biosamplers. DNA is released from the collected SPED and quantified by qPCR.

## MATERIALS AND METHODS

### Synthesis of SPED

SPED synthesis (S1 and S2) was performed in adaption of Paunescu et al.^25^ and characterized regarding DNA load, size and shape. For DNA encapsulation, 4×4ml of silica nanoparticles (110 nm, 50 mg/mL in isopropanol; Pinfire, Frankfurt a. M., Germany) per batch were surface-functionalized in 4 separate falcon tubes by adding 40 µg of N-trimethoxysilylpropyl-N,N,N-trimethylammonium chloride (TMAPS) (50% wt in methanol; abcr, Karlsruhe, Germany) followed by 12 hours of stirring at 900 rotations per minute (rpm) at room temperature. For DNA adsorption to the surface, a 2 mL batch of 150 ng/µL corresponding annealed DNA (sequences see Table 1; Microsynth AG, Balgach, Switzerland) was added to 200 mL ultrapure water (mQ; type 1, 18.2 MΩ·cm at 24°C, Milli-Q®; Merck, Darmstadt, Germany). 0.4 g of the functionalized particles were added to the DNA solution and shaken for 10 seconds. Subsequently, 4 µL TMAPS were added, then the mix was shaken and sonicated for 20 seconds. Next, 62.5 µL of tetraethyl orthosilicate (TEOS) (≥99.0%; Sigma-Aldrich, St. Louis, Missouri, USA) were added, followed by 5 hours of shaking at 600 rpm on a mixer (Vibramax 100; Heidolph, Schwabach, Germany) with a universal clamping attachment (VX 8; IKA, Staufen, Germany). In a further step, 10 mL isopropanol and 5.9 mL TEOS were mixed with 484.1 mL mQ water and combined with the previous mixture. The batch was again stirred at 600 rpm for 4 days, before washing twice with water.

**Table 1:**
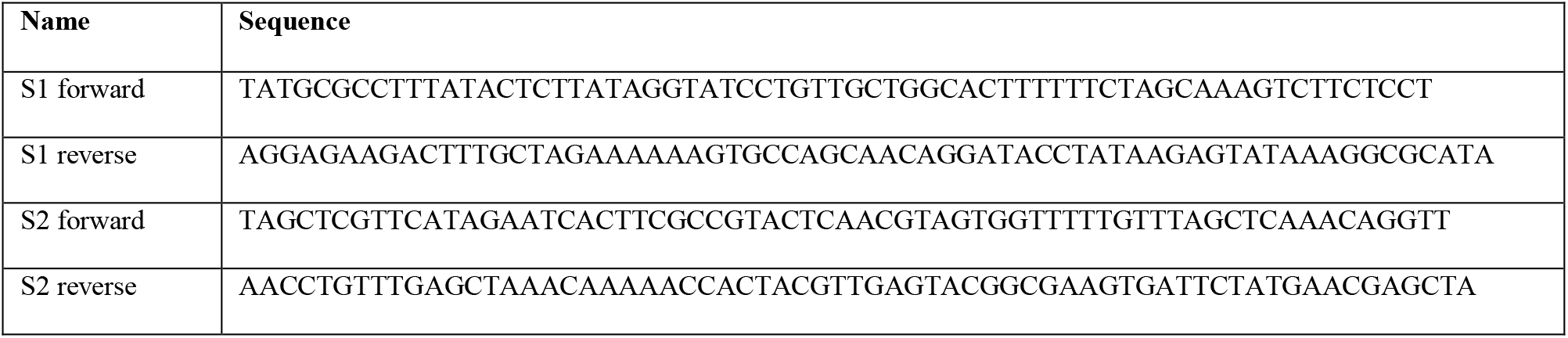
DNA sequences encapsulated in silica for SPED synthesis.

### SPED characterization

DNA load was determined by photospectrometric measurement of DNA concentration in solution before and after encapsulation (NanoDrop 2000c; Thermo Fisher Scientific, Waltham, Massachusetts, US). Particle size distributions of SPED in suspension (prepared in mQ water at ~2 mg/mL) were measured using an analytical photocentrifuge (dispersion analyzer LUMiSizer, light source 470 nm; LUM GmbH,). Transmission profiles were recorded for 2 hours in time intervals of 5 seconds, at a rotational speed of 3000-4000 rpm. Statistical data analysis was performed by SEPView® software (LUM GmbH). Scanning electron microscopy (SEM) was performed on NovaNanoSEM450 device (Field Electron and Ion Company, Hillsboro, Oregon, US) with samples loaded on a TEM grid.

### Aerosolization and droplet characterization

The aerosol experiments were performed in a 277 m^3^ laboratory running at a ventilation throughput of 1840 m^3^/h. A technical layout of the room can be found in the supplementary material, Figure S2d. SPED-containing aerosols were generated by a commercial airbrush gun with a 0.35 mm nozzle (type AFC-101A; Conrad Electronic, Hirschau, Germany) using an air pressure of 1 bar, mounted to a height of 1.6 m on a mobile stand. Phase Doppler anemometry was used to characterize droplet size distribution as created by the airbrush gun used for aerosol generation. The point of measurement was 2 cm in front of the nozzle, using a standard SPED suspension. For a more detailed description refer to the supplementary material, section S1.2.

For each sampling, 5 mL of a 0.2 mg/mL aqueous suspension of the respective SPED species were nebulized over the course of approx. one minute. For the treatment and wash protocol of SPED before dispersion, refer to the supplementary material, section S1.4.

### Aerosol Capture

For aerosol capture, biosamplers, also called flow impingers (Aquaria SRL, Lacchiarella, Italy and SKC Ltd, Dorset, UK), were filled with 20 mL of ultrapure water and equally mounted to a height of 1.6 m. For photographs and descriptions of the different components refer to the supplementary material, Figures S3 and S4. Previous to aerosol dispersion, zero sampling was performed, which consisted of sampling the empty room using the same setup and conditions.

Air samplers were used in combination with 40 L/min piston vacuum pumps (Ningbo Nuolin Mechatronics Co Ltd, Ningbo, China), whereby the air flow rate was reduced to 12.5 L/min using needle valves. If not indicated otherwise, sampling time was 2 hours post-dispersion. For time-resolved experiments, the pumps were briefly stopped at given time points, each time removing 200 µL of the respective capturing liquid from the samples for analysis.

### Quantitative PCR

Immediately after sampling, the collection liquid from each flow impinger was transferred into a 50 mL falcon tube and sonicated for 10 min in an ultrasound bath (model Sonorex Digitec DT 31H; Bandelin, Berlin, Germany) at room temperature. 200 µL were used for further processing and treated with 4 µL buffered oxide etch, consisting of 0.03wt% ammonium hydrogen difluoride (NH_4_FHF, pure; Merck, Darmstadt, USA) and 0.02wt% ammonium fluoride (NH_4_F, puriss.; Sigma-Aldrich, St. Louis, Missouri, USA). After adding the etching reagent, the mix was vortexed and sonicated again. For the subsequent polymerase chain reaction, 5 µL of the broken down SPED solution were mixed with 10 µL of KAPA SYBR FAST qPCR master mix universal (2x; Kapa Biosystems, Wilmington, USA), 1 µL of each primer solution and 3 µL of PCR-grade water (type 1, 18.2 MΩ·cm at 24°C, Milli-Q®; Merck, Darmstadt, Germany). Primer sequences (see Table 2) were ordered in dry state from Microsynth AG (Balgach, Switzerland) and dissolved in PCR-grade water to a working concentration of 10 µM. Samples were run in technical triplicates on a LightCycler® 96 instrument (Roche Molecular Systems, Pleasanton, USA). The qPCR program consisted of a pre-incubation step (240 seconds, 95°C) followed by 40 cycles of 3-step-amplification (2 seconds, 95°C; 12 seconds, 60°C; 4 seconds, 72°C). Cycle values were assigned automatically from qPCR fluorescence curves by the LightCycler® 96 SW 1.1 analysis programme (Roche Molecular Systems, Pleasanton, USA), and conversed to concentrations based on previously measured standard curves. For a more detailed description of the calculations and determination of experimental detection limits refer to supplementary material, sections S1.4-5.

**Table 2:**
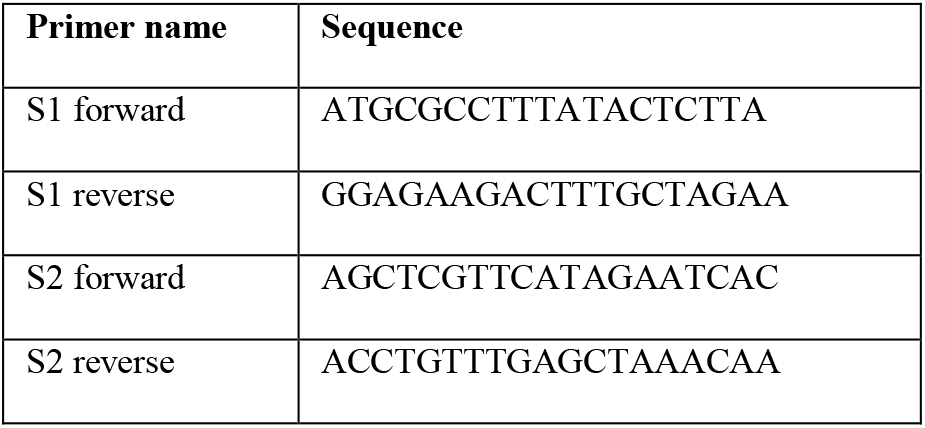
Primer sequences used for qPCR.

### Fog experiment

For visualization of air and ventilation dynamics, fog was generated by a commercial fog machine (Model Rage 600I; BeamZ, Almelo, Netherlands), used in conjunction with the liquid provided with the device (BeamZ fog fluid 1 L High-Density). The fog was dispersed according to instructions by the manufacturer from the same point of origin as the SPED. Per experiment, one load was dispersed, corresponding to 20-30 seconds of continuous operation. Qualitative analysis was performed by eye and documented in images and on video (see supplementary video file).

## RESULTS AND DISCUSSION

Characterization of the two SPED species (S1 and S2) used for the following tracing studies revealed a DNA load of 23 µg and 26 µg dsDNA/mg for S1 and S2 particles, respectively. The median hydrodynamic size was determined to be 133.5 nm for S1 and 160.6 nm for S2, as shown in Figure 2b. This is further confirmed by electron microscopy (Figure 2a), which shows the spherical shape and homogenous size distribution of the particles. The selected core particle size of 110 nm is similar to viruses like adenovirus^46^, influenza A^47^, or SARS-CoV-2^48^, all ranging between 90 and 120 nm in size.

**Figure 2.**
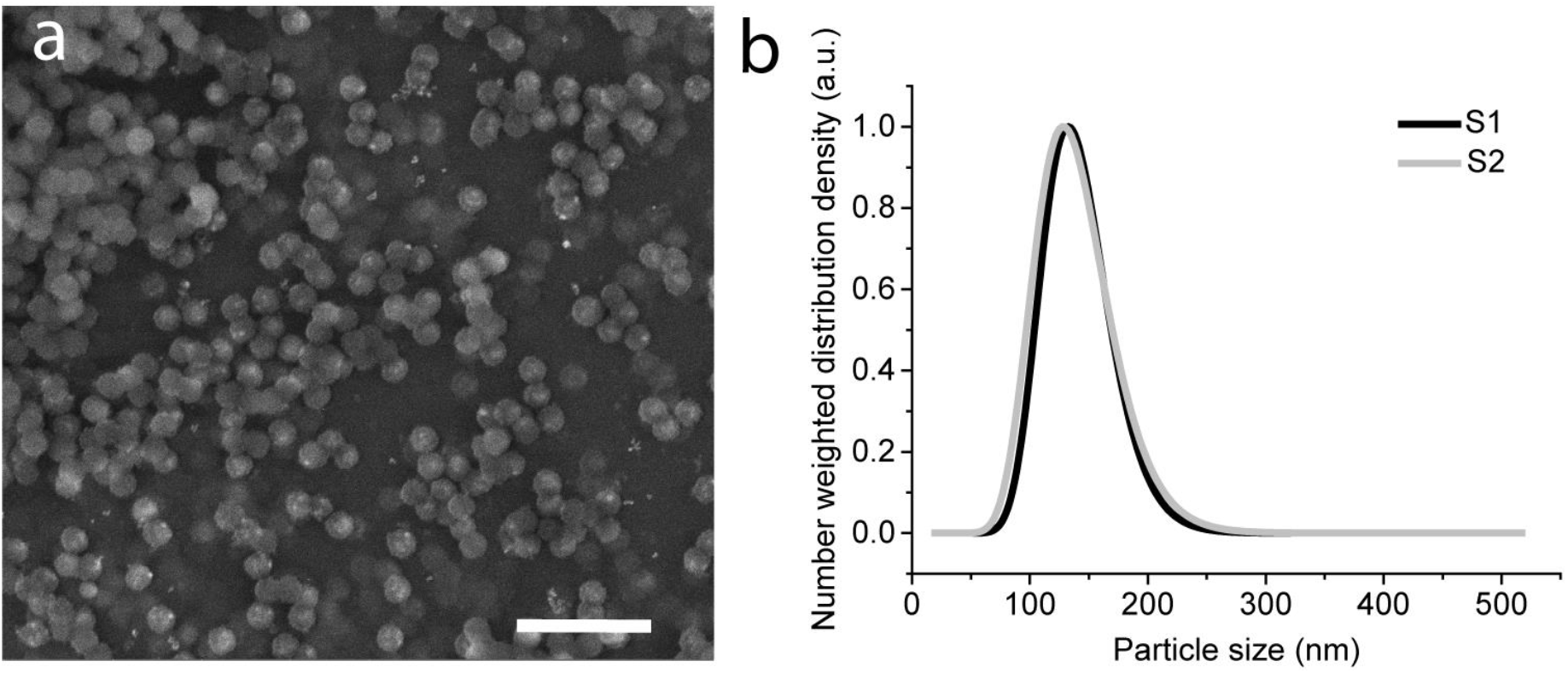
**(a)** Scanning electron microscopy (SEM) image of SPED species S2. Scale bar is 500 nm. **(b)** Hydrodynamic size distribution of two batches of SPED (S1 and S2) by analytical photocentrifugation. For S1, the median is at 133.5 nm, for S2 at 160.6 nm.

In addition to the hydrodynamic diameter of SPED in suspension, the droplet size generated by the airbrush gun was determined using phase Doppler anemometry (PDA), the results of which are shown in the supplementary material, section S1.2.

Figure 3a shows a summary of the setting used to investigate and quantify the distribution of SPED-aerosols indoors. 5 mL of a 0.2 mg/mL particle suspension were dispersed over the time-course of ca. 1 minute in a 277 m^3^ laboratory, running at a ventilation output of 1840 m^3^/h. This corresponds to roughly 6.6 air changes per hour (ACH), or roughly one air change every 10 minutes. For aerosol capture, up to 5 flow impingers were mounted with the inlets at a height of 1.6 m. This height was chosen because it is similar to the level of the nose of an average worker in the laboratory. A graphic depicting the location of the samplers within the test space is shown in Figure 3b. For aerosol capture, each flow impinger was connected to a pump aspirating 12.5 L/min of ambient air, which is close to a human’s minute ventilation for a low-level activity, such as driving a car^49^. Through the focused airflow, aerosols are trapped in the collection water, which can directly be used for further processing and subsequent PCR-analysis. The setup is mobile and only requires electricity from regular sockets and a small compressed air source for the airbrush gun. A battery-powered version of the same setup would also be feasible.

**Figure 3.**
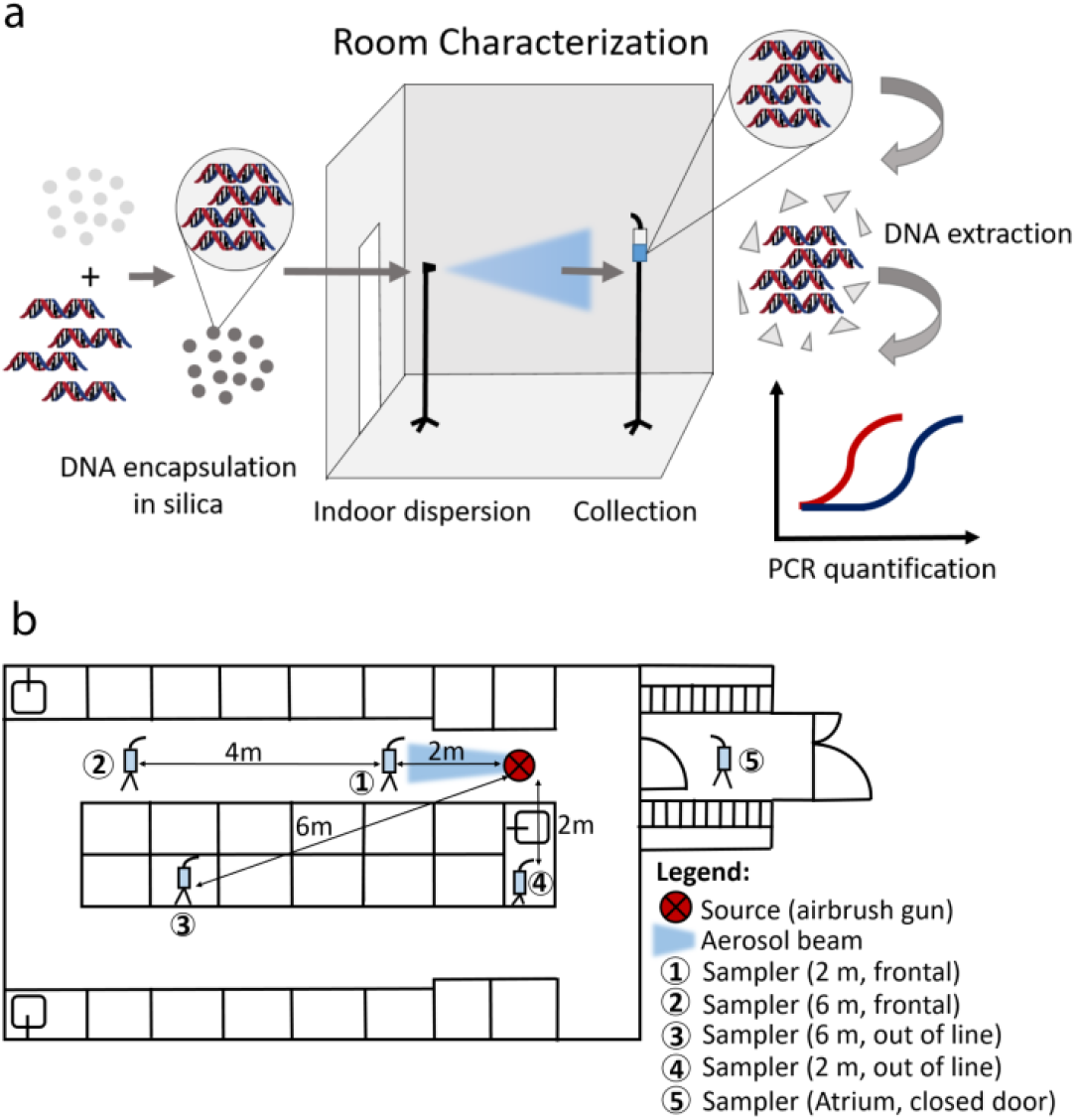
(a): Schematic representation of room characterization experiments. SPED are dispersed in an indoor environment, re-collected using one or several sampling devices and analyzed using quantitative PCR. (b) Room layout of the laboratory used for the experiments, schematically showing benchtops, sinks, fume hoods and hallways. Distances of the five sampling device locations from the source of dispersion are indicated.

As a pilot test, a time-resolved experiment was conducted. For this experiment, two flow impingers were used, mounted 2 m and 6 m frontal to the origin of aerosol flow, respectively, and the test laboratory ran at regular ventilation output. 1 mg of SPED 1 was dispersed and during a continuous sampling time of 120 min, 200 µL samples were removed from the initial 20 mL solution at 14 time points and analyzed separately for their SPED concentration. Figure 4a shows the integrated aerosol levels measured in the two impingers over time. The first measurement, at t=0, was taken before dispersion and is thus the experimental negative control, marking the minimal limit of detection (MLD) of 4.5·10^−9^ mg/mL in the sampling solution for this experiment. Assuming a conservative physical collection efficiency for the impingers of 10%, as estimated from literature data^50–52^, this corresponds to a particle concentration detection limit of 6·10^−2^ ng/m^3^ of sampled air. Comparing this value to commercial monitoring systems for inorganic particulate matter 2.5 (PM_2.5_)^53, 54^ the MLD of the present method is at least one order of magnitude lower. More information on the impinger efficiency, concentration measurements and how detection limits were determined can be found in the supplementary material, section S1.5.

**Figure 4:**
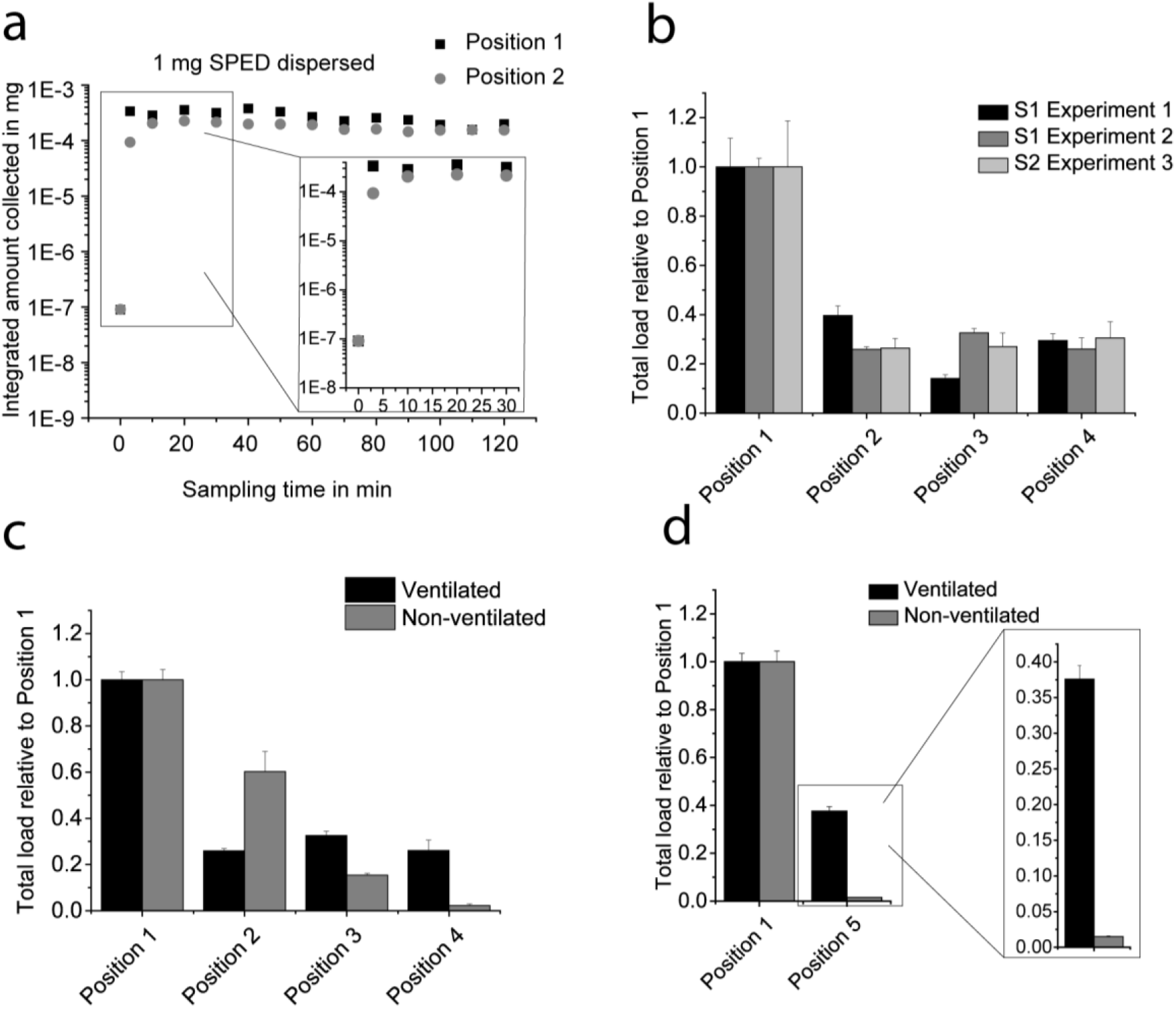
Time- position- and ventilation-dependent effects as measured by air-sampling after dispersion of SPED aerosols. Error bars are calculated from standard deviations of PCR triplicates. Position numbers refer to sampler location in the room as depicted in Figure 3b. (**a)** Time-dependent measurement using two air samplers. The y-axis indicates the integrated amount of collected SPED (in mg) over a total time of 120 min. The measurement point at 0 min was taken immediately before dispersion as a zero control. The first five measurements are zoomed in for clarity. **(b)** Measurement of position-dependent relative exposure of 4 impingers. Two experiments were performed with particles S1 and a third using particles S2. The values reflect the total amount of particles collected after a two hour sampling time. The data are normalized to the first position in each set. **(c)** Total load relative to position 1 per impinger with (data from S1 experiment 2) and without ventilation under equal conditions as in (c). **(d)** Same as (c), but comparing position 1 to an additional position in the adjacent atrium (position 5), which is additionally zoomed in for clarity.

Following the aerosol generation, the amount of SPED collected rises sharply in both impingers, prior to reaching a plateau load in the range of 3·10^−4^ mg and 2·10^−4^ mg, respectively. At position 1, the maximum is reached within the first measurement increment after 3 minutes, whereas at a distance of 6 m, there is a distinguishable saturation curve reaching the plateau after an estimated 10-20 minutes, attributing to the fact that the aerosols reach the second sampling point with a delay due to further traveling distance. This position-dependent effect is also reflected in the total collection loads, which correspond to estimated position-specific dosages of 0.35% and 0.2% of the total amount of released SPED. Throughout the measurement time, there are some fluctuations; however, these deviations are within the expected range for PCR reactions, as discussed further below.

To further investigate position-dependency, the total load over 120 min was measured under otherwise equal sampling conditions. In addition to the two impingers used in the previous experiments, two locations with a 2 m offset relative to the airbrush gun’s line of flight were sampled simultaneously (labelled positions 3 and 4 in Figure 3b). The results of three separate runs – two with barcode S1 and one with S2 - are displayed in Figure 4b. To relate total load to the distance from the source, the data are always normalized to the 2 m frontal position, which is standing within the aerosol flow and therefore considered an experimental positive control. The data show that relative exposure is much lower at all three locations that are either further apart from the source, and/or not in the direct line of flight. Additionally, the three experiments qualitatively compare to each other, independent of the DNA barcode used.

In a next step we wanted to determine the influence of ventilation to aerosol distribution and clearance. We therefore repeated the previous measurements, but turned off the ventilation system with doors and windows remaining closed, as before. Figure 4c compares total load after 2 hours of sampling at the four previous sampling points with ventilation on and off, respectively, again relative to the position close to the point of aerosolization. This experiment revealed a more pronounced concentration difference between the individual sampling points. This result follows intuition, as in the non-ventilated scenario the air flow in the room is more stagnant as opposed to the forced convection found in the experiments under room ventilation.

To assess whether SPED could also be detected in an adjacent room, we placed an additional sampler in the atrium next to the test laboratory (position 5 from Figure 3b), separated by a closed door, with a cubature of 30 m^3^. These results are displayed separately in Figure 4d, comparing position 5 to position 1. The expectation was that exposure in the atrium would only be minimal. Interestingly though, the exposure is considerable in the ventilated scenario, but very low without ventilation. Based on this outcome, we hypothesized that ventilation dynamics could lead to a suction effect from the main room to the atrium. The layout of the ventilation piping (see supplementary material, Figure S2d), which shows a connection between the two rooms, is compatible with this theory.

As these results were unexpected, we wanted to reconfirm them using another method. We therefore used a fog machine to visualize air dynamics and to add qualitative evidence to the previous results. The fog was generated at the same position as the previous source and a video camera was installed in the atrium. Indeed, with ventilation running, fog could be visually detected with a delay of ca. 5 min (see supplementary video file) seemingly coming from the ventilation pipe. The fog does not allow for quantification or localized detection and has a comparatively high detection limit, but it was sufficient to qualitatively confirm our measurements. This shows that the method introduced here is suitable to detect unexpected effects in the distribution dynamics of aerosols in a real-world setting, as caused by a specific ventilation system. There are indications that ventilation systems and air-conditioning can contribute to the spreading of airborne diseases instead of preventing it^10^, which is why it is already recommended to replace air recirculation by increased inflow of outdoor air^55^. The results show that the described method is able to measure such effects and could for example be employed in finding appropriate ventilation settings to limit aerosol spread.

The present proof-of-concept shows that a simple setup can be used to reliably measure time-, position-, and ventilation-dependent relative aerosol loads indoors using different DNA barcodes. Current limitations of the sampling and detection method are a direct result of PCR analysis, which requires normalization to achieve quantitative results. Furthermore, PCR data are logarithmic to the concentration levels. Consequently, result quantification requires a range of control experiments, and small concentration differences are more difficult to detect, requiring numerous sample replicas. Furthermore, the individual measurements were conducted in a real-world environment under similar conditions, but without perfect control of external parameters. Factors such as relative humidity, temperature and the presence and concentration of other particulate matter in the ambient air can influence aerosol dynamics and lead to a difference in absolute aerosol levels accessible to measurement.

In spite of the limitations mentioned above, the SPED-based method introduced here has several important benefits, the combination of which makes it novel and unique among aerosol tracers. Important features are the versatility, sensitivity and specificity towards the sampled barcode and suitability to detect effects related to air circulation. The silica layer protects the DNA from physical and chemical damage^25^, which is an additional advantage over other DNA-based tracers and makes SPED suitable for indoor and outdoor use alike. The costs of SPED are estimated at 500 USD/g, which benchmarks the price of particles for a single experiment of the scale discussed here at less than 1 USD.

Even though flow impingers were used in this experimental setup, the use of SPED in combination with other means of aerosol capturing, including surface sampling^36^, is conceivable. And while the airbrush gun used in this pilot study presents a cost-effective, user-friendly solution, SPED could equally be used in combination with advanced dispersion systems to e.g. mimic human breathing or coughing^56^. Furthermore, it has been shown that SPED detection is possible at a single particle level in solution^57^. Thus, when limiting barcodes to single use and with a further focus on optimization of the respective sampling and measurement conditions, an additional improvement of the detection limit in the range one or two orders of magnitude is conceivable. Consequently, the flexible barcodes offer countless combination possibilities in future measurements, such as simultaneous surface and air sampling with multiple sources and detectors at extremely low detection limits.

## CONCLUSIONS

In conclusion, this study presents a simple, cost-effective setup for investigating aerosol distribution using a novel tracing agent. The SPED-based platform offers a foundation for a range of real-world tracing scenarios at low detection limits using qPCR as method of analysis. Future applications are the study of aerosol flow in complicated architectural settings, as well as the dynamics of free convection resulting from heating, daytime effects and door/window arrangements. In the near future, SPED could additionally be tuned in size, made biodegradable, or be designed to mimic specific pathogens or hazardous pollutants. These are ideal prerequisites for developing a robust platform to examine places of interest regarding potential health hazards and environmental risks.

## Data Availability

Raw data are available from authors upon request.

## AUTHOR CONTRIBUTIONS

A.M.L. performed experiments and wrote the first manuscript draft, J.K. prepared and analyzed particles and assisted in exposure experiments. R.N.G. supervised the project, R.N.G. and W.J.S. conceived the project. The manuscript was written through contributions of all authors. All authors have given approval to the final version of the manuscript.

## ACKNOWLEDGEMENTS

We thank Prof. Ulrike Lohmann and Jörg Wieder for supplying us with additional biosampling devices, and Nikita Kobert for providing the laboratory space and technical support. This work was financially supported by ETH Zürich.

## Notes

### Competing Interest Statement

The authors declare the following competing financial interest: R.N.G. and W.J.S. declare a financial interest in the form of IP on DNA encapsulation licensed to Haelixa AG, of which R.N.G. and W.J.S. are shareholders. A.M.L. and J.K. have no competing financial interests.

### Funding Statement

This work was financially supported by ETH Zurich.

### Author Declarations

Since no research with humans was conducted and no personal data were collected or used for this study, no ethics approval was necessary. The research project was approved based on a risk analysis by the Safety, Security, Health and Environment administrative department of ETH Zurich.

