## Supplementary Material for "Silica-encapsulated DNA tracers for measuring aerosol distribution dynamics in real-world settings"

### Contents

S1 Extended Materials and Methods ... 2
**S1.1 Barcode design and DNA annealing** ... 2
**S1.2 Phase Doppler Anemometry** ... 2
**S1.3 Air sampling** ... 4
**S1.4 Conversion of qPCR data to collected aerosol load** ... 6
**S1.5 Conversion of collected load to aerosol concentration in sampled air and experimental detection limits** ... 9
S2 Raw Data of air-sampling experiments ... 10
**S2.1 Time-resolved experiment** ... 10
**S2.2 Air-sampling experiments with ventilation** ... 11
**S2.2.1 Experiment 1, Particles S1** ... 11
**S2.2.2 Experiment 2, Particles S1** ... 12
**S2.2.3 Experiment 3, Particles S2** ... 13
**S2.2.4 Air-sampling experiment without ventilation, Particles S1** ... 14

### S1 Extended Materials and Methods

#### S1.1 Barcode design and DNA annealing

DNA strands were designed at a length of 65 nucleotides with a GC-content of 40%. The sequences were ordered as single strands in dried state from Microsynth AG (Balgach, Switzerland). Previous to particle loading, the forward and reverse strands were dissolved to a final concentration of 5 g/L in annealing buffer (Tris 10 mM pH 7.5-8.0, EDTA 1 mM, NaCl 50 mM), mixed together in equal parts and annealed for 5 min at 95 °C before cooling slowly to room temperature.

#### S1.2 Phase Doppler Anemometry

In addition to the hydrodynamic volume of SPED in suspension, the droplet size generated by the airbrush gun was determined using phase Doppler anemometry (PDA). PDA was measured on a custom-made device at ETH Zurich. To measure droplet size distribution, pure water was used, at a flow rate of ca. 2 mL/min resulting from an air pressure of 2 bar from a mobile source. The point of measurement was 2 cm in front of the nozzle.

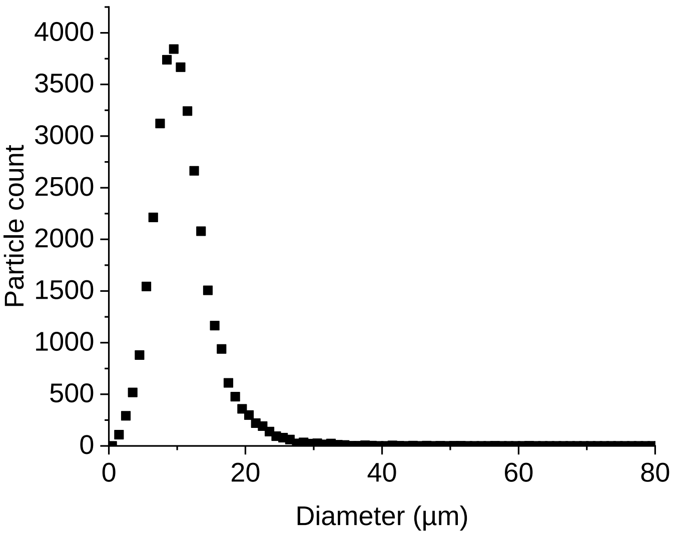


Figure S1: Droplet size distribution as measured by phase Doppler anemometry.

The droplet size distribution is shown in Figure S1. The measurement revealed an arithmetic mean droplet size of 10.8 ± 4.4 µm. There is abundant literature characterizing size distributions of droplets as created by human sneezing, coughing, speaking and breathing. A comparison between methods yielded a very broad range of observed sizes, with most of the studies taken into account reporting values between 150 nm and 180 µm for the majority of droplets[1]. These numbers ultimately depend on the method of impaction and detection, point of measurement and overall setting. Generating an experimental setup that mimics human respiration was not a primary focus of this study. Nonetheless, the droplet size distribution created by the airbrush gun seems to be within in the previously observed range for respiratory droplets. Additionally, according to the Wells evaporation-falling curve, droplets in the observed size range will, under typical conditions observed indoors, rapidly dry out to form droplet nuclei, which can remain suspended in air for significant time periods and contribute to airborne disease transmission over distances beyond close proximity to the source.[2, 3]

##### S1.3 Air sampling

All field tests were performed in the same laboratory with an area of ca. 80 $m^2$ and a cubature of ca. 277 $m^3$ and a maximal ventilation power of 9200 $m^3/h$. The room is partitioned in two sides by a double-row of workbenches (see Figure S2).

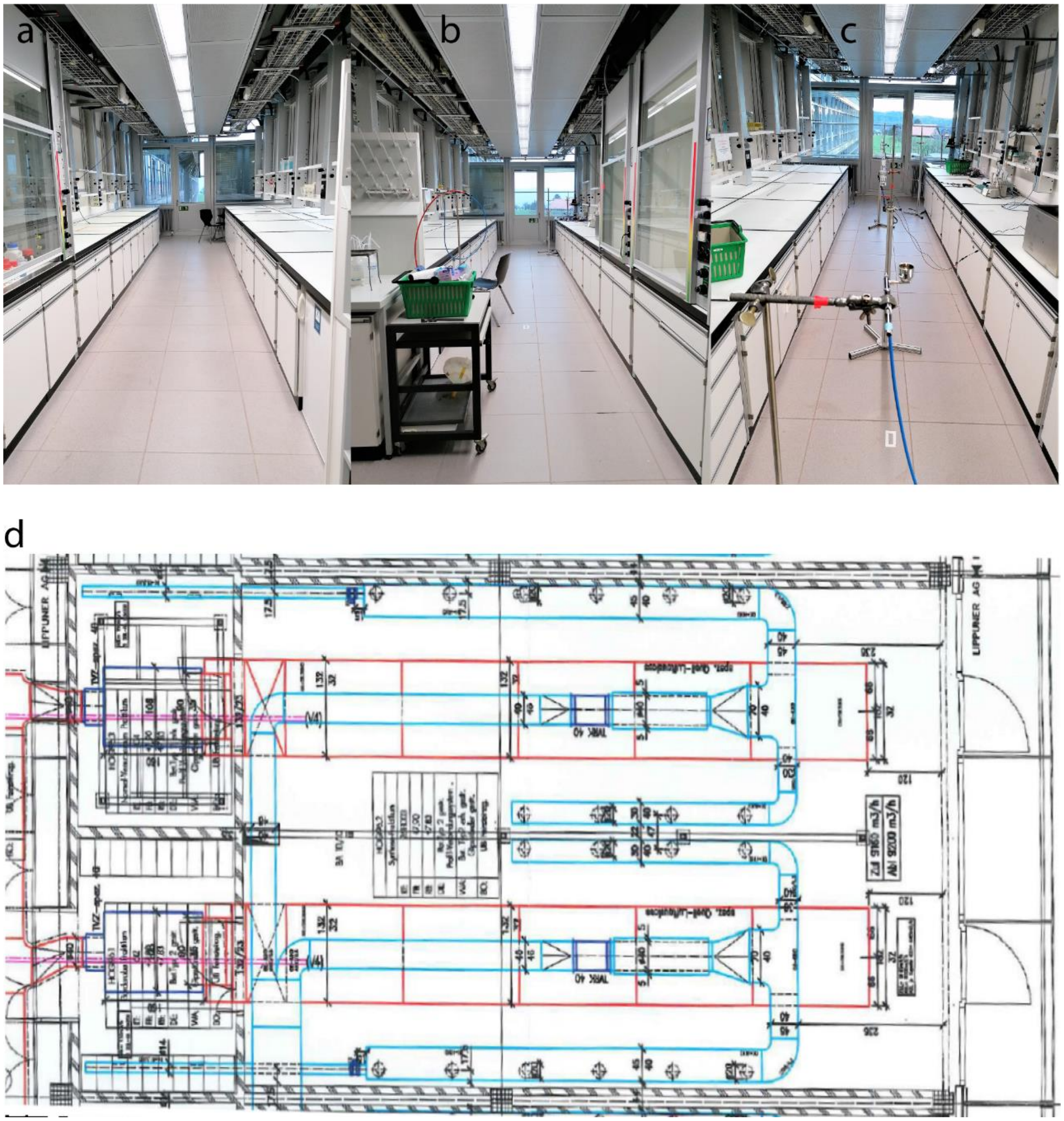


Figure S2: Partitioned laboratory used for field testing. (a) Left side of the room. (b) Right side of the room. (c) Right side of the room including experimental setup, photographed from the angle of aerosol dispersion with the airbrush gun in the front of the frame. (d) Room layout of the laboratory, rotated 90 degrees clockwise. Red lines indicate supply air piping, blue lines exhaust air piping.

Droplets were generated using an airbrush gun as shown in Figure S3.

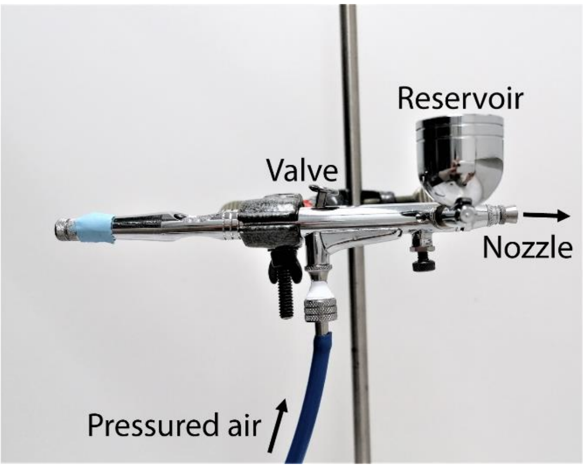


Figure S3: Airbrush gun setup.

The resulting aerosols were re-captured for analysis using Biosampler Flow Impingers, as shown in **Error! Reference source not found.**.

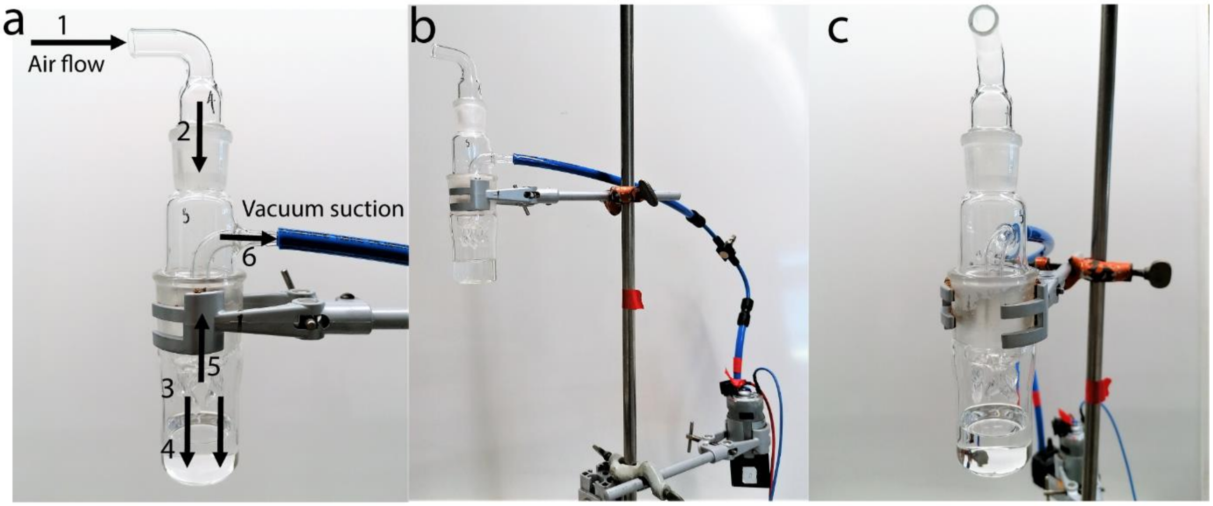


Figure S4: Flow impinger setup. (a) Side view of mounted impinger. Arrows indicate direction of airflow. A vacuum pump draws in air through the impinger at a rate of 12.5 l/min. The attracted air enters the sampling device at the inlet (1). It then travels down the body (2) into small outlet branches (3), which leads to highly pressured air meeting the water (4). The air then "bubbles" through the liquid, where aerosols are captured while the air is sucked back up the central tube (5), leaving the device through the outlet (6) through the tubing, at the end of which the pump is attached (not shown in this frame). (b) shows the fully mounted setup, with the flow impinger at the top left, which is connected to the vacuum pump (bottom right) through PVC tubing (in blue). The tubing contains a needle valve, through which the flow rate of the pump is controlled. (c) Front view of the whole setup.

#### S1.4 Conversion of qPCR data to collected aerosol load

Dilution series of the SPED species used for the experiments were measured to relate known particle concentrations with cycle values from qPCR measurements. 5-6 concentrations (diluted in powers of ten) were measured under equal qPCR conditions. Stocks were prepared by washing 1 to 2 mL of SPED twice to remove any leached DNA before measuring calibration data and using the respective aliquot for field experiments. Washing was performed by centrifugation (5 min, 13'000 rpm), followed by removal of 95% of the water volume before re-suspending the pellet in fresh PCR-grade water. For each aliquot of washed SPED, a separate calibration curve was measured, which was then used for all experiments using the same stock. The qPCR cycle values were plotted against the logarithmic known particle concentration (in mg/mL). Linear regression of the single log chart yields a curve in the form of $y = ax + b$, where y is the cycle value and x is the log (or ln) of the concentration, therefore concentration $c = e^{y/m-b}$. Tables S1-S3 show the PCR data of the standard dilution series, Table S4 lists the regression parameters calculated from said data, with the corresponding graphs shown in Figure S5.

Table S1: PCR data from dilution series of S1 Stock 1

| Conc. in g/L | Cq av | STD |
|---|---|---|
| **1.00E-03** | 5.62 | 0.15 |
| **1.00E-04** | 9.06 | 0.15 |
| **1.00E-05** | 12.92 | 0.07 |
| **1.00E-06** | 16.61 | 0.22 |
| **1.00E-07** | 20.78 | 0.64 |

**Conc. in g/L**: SPED concentration in suspension

**Cq av**: Mean of Cq-values of PCR triplicates

**Cq STD:** Standard deviation of PCR triplicates

Table S2: PCR data from dilution series of S1 Stock 2

| Conc. in g/L | Cq av | Cq STD |
|---|---|---|
| **1.00E-03** | 5.86 | 0.21 |
| **1.00E-04** | 9.71 | 0.05 |
| **1.00E-05** | 13.61 | 0.15 |
| **1.00E-06** | 17.21 | 0.18 |
| **1.00E-07** | 20.7 | 0.3 |
| **1.00E-08** | 24.37 | 0.59 |

**Conc. in g/L**: SPED concentration in suspension

**Cq av**: Mean of Cq-values of PCR triplicates as calculated by LightCycler96 Software

**Cq STD:** Standard deviation of PCR triplicates as calculated by LightCycler96 Software

Table S3: PCR data from dilution series of S2 Stock 1

| Conc. in g/L | Cq av | Cq STD |
|---|---|---|
| **1.00E-03** | 6.56 | 0.13 |
| **1.00E-04** | 10.88 | 0.17 |
| **1.00E-05** | 14.47 | 0.17 |
| **1.00E-06** | 19.26 | 0.14 |
| **1.00E-07** | 23.83 | 0.06 |
| **1.00E-08** | 27.69 | 0.17 |

**Conc. in g/L**: SPED concentration in suspension

**Cq av**: Mean of Cq-values of PCR triplicates as calculated by LightCycler96 Software

**Cq STD:** Standard deviation of PCR triplicates as calculated by LightCycler96 Software

Table S4: Regression parameters for concentration determination. S1 stock 1 was used for all S1 experiments in the ventilated setting, S1 stock 2 for the non-ventilated setting. S2 stock 1 was used for all experiments involving S2 SPED.

| **Batch** | **a** | **b** | **$R^2$ (coefficient of determination)** |
|---|---|---|---|
| S1 stock 1 | -1.645 | -5.937 | 0.999 |
| S1 stock 2 | -1.602 | -5.047 | 0.999 |
| S2 stock 1 | -1.852 | -6.345 | 0.999 |

To determine total load, PCR cycle values were conversed to concentrations using the above formula, then multiplied with the volume of remaining collection liquid after sampling (thereby accounting for volume loss by evaporation and distribution within the setup beyond the sampling container). For time-resolved experiments, it was estimated that volume loss occurs linearly throughout the sampling time. Therefore, the volume at each time point was assumed to have reduced linearly from the beginning of measurement, in addition to the reduction caused by removing 200 µL per sample. Error bars were calculated based on the standard deviation of technical triplicates in qPCR data.

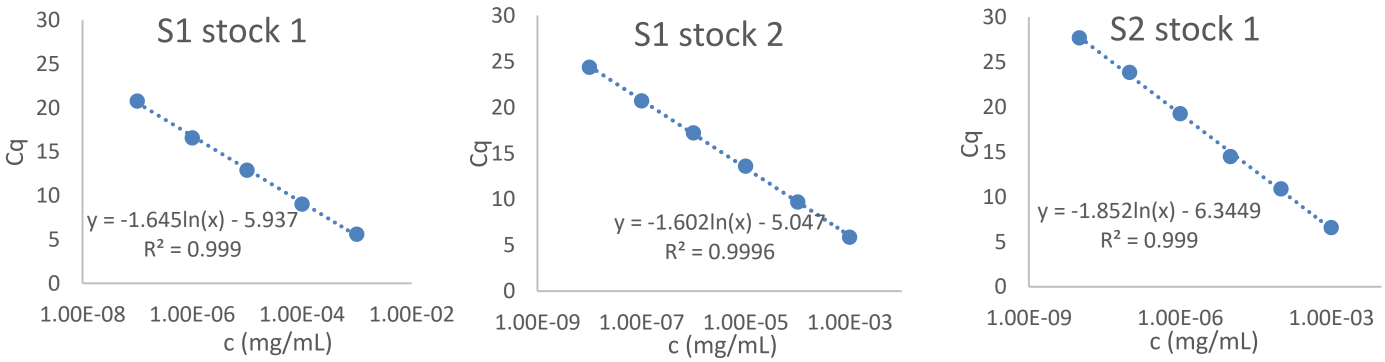


Figure S5: Regression curves of SPED stocks used for the expermients. (a) S1 stock 1, (b) S2 stock 2, (c) S2 stock 1.

#### S1.5 Conversion of collected load to aerosol concentration in sampled air and experimental detection limits

The zero sampling measuring the highest background concentration was considered as minimal limit of detection (MLD) for a given experiment. This is a conservative approach to account for potential (cross-)contamination from experimental processing in the laboratory. To calculate the detection limit, the total load as calculated from the average Cq-value from technical triplicates (see chapter “Conversion of qPCR data to aerosol load”) was divided by the volume of air sampled during the experiment (corresponding to 1.5 $m^3$ air for a 120 min experiment with a sampling rate of 12.5 L/min) and by the conservatively estimated collection efficiency of 10%. The collection efficiency of the biosamplers was roughly estimated based on literature findings. For the used type of sampling device, Willeke et al.[4] found a collection efficiency of ca. 80% for polystyrene latex particles of 300 nm diameter and a 15% decrease in overall efficiency for 2 h sampling in water. Li et al.[5] reported a physical collection efficiency of below 60% for 150 nm particles, but used a smaller scale of biosampler with less collection liquid in their experiment. Hogan et al.[6] in contrast found an efficiency of of only roughly 12-14% for the same size of particles, as generated from atomized bacteriophage suspensions. We decided to take the most prudent approach, combining the lowest of these reported collection efficiencies with the abovementioned loss of efficiency over 2 hours of sampling. This leaves an estimated value for collection efficiency of ca. 10%. This conservative and very rough approximation is likely to lead to an overestimation of the actual detection limit, i.e. in reality this would correspond to even lower concentrations.

### S2 Raw Data of air-sampling experiments

#### S2.1 Time-resolved experiment

Table S5: qPCR data from time-resolved experiment, using S1 stock 1

| Time in min | Cq Position 1 | Cq Position 2 | STD 1 | STD 2 |
|---:|---:|---:|---:|---:|
| 0 | 29.25 | 29.23 | 0.39 | 0.31 |
| 3 | 13.98 | 16.36 | 0.11 | 0.08 |
| 10 | 14.24 | 14.86 | 0.16 | 0.03 |
| 20 | 13.77 | 14.62 | 0.1 | 0.3 |
| 30 | 13.93 | 14.65 | 0.07 | 0.03 |
| 40 | 13.53 | 14.73 | 0.19 | 0.03 |
| 50 | 13.71 | 14.67 | 0.08 | 0.2 |
| 60 | 14.03 | 14.65 | 0.08 | 0.25 |
| 70 | 14.27 | 14.93 | 0.02 | 0.13 |
| 80 | 13.97 | 14.83 | 0.21 | 0.09 |
| 90 | 14.04 | 14.95 | 0.01 | 0.04 |
| 100 | 14.32 | 14.75 | 0.17 | 0.12 |
| 110 | 14.62 | 14.64 | 0.07 | 0.16 |
| 120 | 14.1 | 14.57 | 0.16 | 0.08 |

1 mg SPED was dispersed. At each timepoint, 200 µL sample were removed from the collection liquid.

**Time in min:** Integrated sampling time after SPED dispersion (0 min is experimental negative control before dispersion and determines the detection limit).

**Cq Position 1:** Average Cq-value (quantification cycle) as calculated by Lightcycler96 Software from PCR triplicates at a given timepoint at measurement position 1 (2 m frontal to point of dispersion).

**Cq Position 2:** Average Cq-value (quantification cycle) as calculated by Lightcycler96 Software from PCR triplicates at a given timepoint at measurement position 2 (6 m frontal to point of dispersion).

**STD 1:** Standard deviation of PCR triplicates from measurement position 1

**STD 2:** Standard deviation of PCR triplicates from measurement position 2

Volume of collection liquid left after 2h: 12.5 mL (the loss of 7.5 mL was assumed to occur at a constant rate over time).

#### S2.2 Air-sampling experiments with ventilation

##### S2.2.1 Experiment 1, Particles S1

Table S6: qPCR data from air-sampling experiment 1, using particles S1 stock 1

| Position | Cq mean | Cq STD | Final volume (mL) |
|---|---|---|---|
| 1 | 10.89 | 0.13 | 15 |
| 2 | 12.24 | 0.09 | 13.5 |
| 3 | 14.11 | 0.1 | 15 |
| 4 | 12.9 | 0.07 | 15 |
| 5 | 12.56 | 0.04 | 12 |
| Zero 1 | 28.77 | 0.19 | 14 |
| Zero 2 | 31.87 | 0.76 | 14 |
| Zero 3 | 31.89 | 2.86 | 15 |
| Zero 4 | 32.26 | 2.48 | 15 |
| Zero 5 | 29.75 | 0.28 | 12.5 |

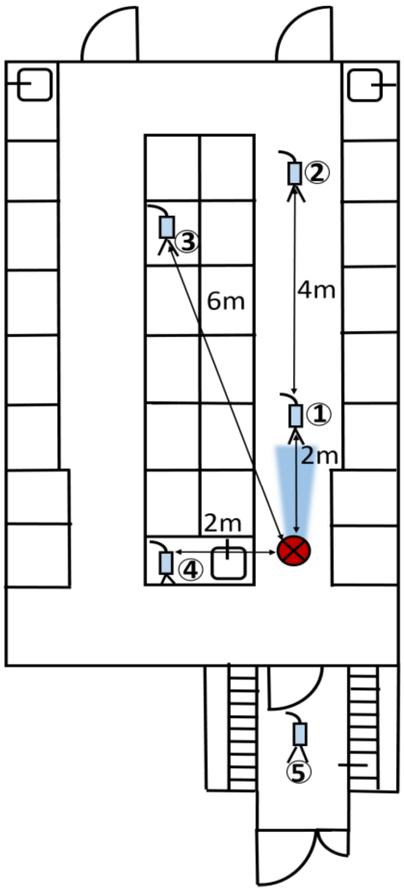


Figure S6: Room layout with indicated sampling positions.

1 mg SPED was dispersed before sampling for 2 h (120 min).

**Position:** Refers to spatial position of the respective collection device, as indicated in Figure S6. “Zero” refers to sampling at the respective position before dispersion of SPED (experimental negative control). The lowest zero Cq value (corresponding to the highest concentration) was used to calculate the experimental detection limit.

**Cq mean:** Average Cq-value (quantification cycle) as calculated by Lightcycler96 Software from PCR triplicates at a given measurement position.

**Cq STD:** Standard deviation of Cq-values of PCR triplicates at a given measurement position

**Final volume:** Volume of collection liquid left in the collector from the original 20 mL after 2h of continuous sampling.

##### S2.2.2 Experiment 2, Particles S1

Table S7: qPCR data from air-sampling experiment 2, using particles S1 stock 1

| Position | Cq mean | Cq STD | Final volume (mL) |
|---|---|---|---|
| 1 | 11.54 | 0.04 | 15 |
| 2 | 13.76 | 0.05 | 15 |
| 3 | 13.21 | 0.08 | 13.5 |
| 4 | 13.75 | 0.26 | 15 |
| 5 | 12.85 | 0.07 | 12.5 |
| Zero 1 | 32.96 | 0.47 | 13 |
| Zero 2 | 33.4 | 0.57 | 14 |
| Zero 3 | 33.69 | 0.9 | 15 |
| Zero 4 | 30.67 | 2.13 | 15 |
| Zero 5 | 33.7 | 0.25 | 12 |

1 mg SPED was dispersed before sampling for 2 h (120 min).

**Position:** Refers to spatial position of the respective collection device, as indicated in Figure S6. “Zero” refers to sampling at the respective position before dispersion of SPED (experimental negative control). The lowest zero Cq value (corresponding to the highest concentration) was used to calculate the experimental detection limit.

**Cq mean:** Average Cq-value (quantification cycle) as calculated by Lightcycler96 Software from PCR triplicates at a given position.

**Cq STD:** Standard deviation of Cq-values of PCR triplicates at a given measurement position

**Final volume:** Volume of collection liquid left in the collector from the original 20 mL after 2h of continuous sampling.

##### S2.2.3 Experiment 3, Particles S2

Table S8: qPCR data from air-sampling experiment 3, using particles S1, stock 2

| Position | Cq mean | Cq STD | Final volume (mL) |
|---|---|---|---|
| 1 | 15.69 | 0.23 | 13 |
| 2 | 17.94 | 0.13 | 10 |
| 3 | 18.52 | 0.27 | 14 |
| 4 | 18.42 | 0.29 | 15 |
| 5 | 16.79 | 0.08 | 13 |
| Zero 1 | 29.81 | 0.22 | 12.5 |
| Zero 2 | 28.94 | 0.26 | 13 |
| Zero 3 | 29.22 | 0.23 | 15 |
| Zero 4 | 29.51 | 0.18 | 15 |
| Zero 5 | 29.62 | 0.43 | 12 |

1 mg SPED was dispersed before sampling for 2 h (120 min).

**Position:** Refers to spatial position of the respective collection device, as indicated in Figure S6. "Zero" refers to sampling at the respective position before dispersion of SPED (experimental negative control). The lowest zero Cq value (corresponding to the highest concentration) was used to calculate the experimental detection limit.

**Cq mean:** Average Cq-value (quantification cycle) as calculated by Lightcycler96 Software from PCR triplicates at a given measurement position.

**Cq STD:** Standard deviation of Cq-values of PCR triplicates at a given measurement position

**Final volume:** Volume of collection liquid left in the collector from the original 20 mL after 2h of continuous sampling.

##### S2.2.4 Air-sampling experiment without ventilation, Particles S1

Table S9: qPCR data from air-sampling experiment without ventilation, using particles S1, stock 1

| Position | Cq mean | Cq STD | Final volume (mL) |
|---|---|---|---|
| 1 | 12.72 | 0.05 | 12 |
| 2 | 13.07 | 0.21 | 9 |
| 3 | 15.84 | 0.06 | 13 |
| 4 | 18.98 | 0.43 | 13.5 |
| 5 | 19.68 | 0.06 | 14 |
| Zero 1 | 23.34 | 0.11 | 11 |
| Zero 2 | 21.63 | 0.05 | 10 |
| Zero 3 | 27.26 | 0.3 | 14 |
| Zero 4 | 26.45 | 0.19 | 13 |
| Zero 5 | 31.62 | 2.19 | 12 |

1 mg SPED was dispersed before sampling for 2 h (120 min).

**Position:** Refers to spatial position of the respective collection device, as indicated in Figure S6. “Zero” refers to sampling at the respective position before dispersion of SPED (experimental negative control). The lowest zero Cq value (corresponding to the highest concentration) was used to calculate the experimental detection limit.

**Cq mean:** Average Cq-value (quantification cycle) as calculated by Lightcycler96 Software from PCR triplicates at a given measurement position.

**Cq STD:** Standard deviation of Cq-values of PCR triplicates at a given measurement position

**Final volume:** Volume of collection liquid left in the collector from the original 20 mL after 2h of continuous sampling.
